# Combined near infrared photoacoustic imaging and ultrasound detects vulnerable atherosclerotic plaque

**DOI:** 10.1101/2023.06.11.23291099

**Authors:** Martin Karl Schneider, James Wang, Aris Kare, Shaunak S. Adkar, Darren Salmi, Caitlin F. Bell, Tom Alsaigh, Dhananjay Wagh, John Coller, Aaron Mayer, Sarah J. Snyder, Alexander D. Borowsky, Steven R. Long, Maarten G. Lansberg, Gary K. Steinberg, Jeremy J. Heit, Nicholas J. Leeper, Katherine W. Ferrara

**Affiliations:** Molecular Imaging Program at Stanford and Bio-X Program, Department of Radiology, Stanford University School of Medicine, Palo Alto, CA 94305, USA; Department of Surgery, Division of Vascular Surgery, Stanford University School of Medicine, Stanford, CA 94305, USA; Department of Pathology, Stanford University School of Medicine, Palo Alto, CA 94305, USA; Sequencing Group Stanford Genomics, Stanford University School of Medicine, Palo Alto, CA 94305, USA; Enable Medicine, Menlo Park, CA, 94025, USA; Department of Radiology and Neurosurgery, Stanford University School of Medicine, Palo Alto, CA 94305, USA; Department of Pathology and Laboratory Medicine, UC Davis School of Medicine, Davis, CA 95616, USA; Department of Neurosurgery, Stanford University School of Medicine, Palo Alto, CA 94305, USA; Department of Pathology, University of California San Francisco, San Francisco, CA 94110, USA; Department of Neurology and Neurological Sciences, Stanford University School of Medicine, Palo Alto, CA 94305, USA

**Keywords:** Photoacoustic imaging, spatial transcriptomics, atherosclerosis, vulnerable plaque, NIR biomarker

## Abstract

Atherosclerosis is an inflammatory process resulting in the deposition of cholesterol and cellular debris, narrowing of the vessel lumen and clot formation. Characterization of the morphology and vulnerability of the lesion is essential for effective clinical management. Photoacoustic imaging has sufficient penetration and sensitivity to map and characterize human atherosclerotic plaque. Here, near infrared photoacoustic imaging is shown to detect plaque components and, when combined with ultrasound imaging, to differentiate stable and vulnerable plaque. In an *ex vivo* study of photoacoustic imaging of excised plaque from 25 patients, 88.2% sensitivity and 71.4% specificity were achieved using a clinically-relevant protocol. In order to determine the origin of the near-infrared auto-photoacoustic (NIRAPA) signal, immunohistochemistry, spatial transcriptomics and proteomics were applied to adjacent sections of the plaque. The highest NIRAPA signal was spatially correlated with bilirubin and associated blood-based residue and inflammatory macrophages bearing CD74, HLA-DR, CD14 and CD163 markers. In summary, we establish the potential to apply the NIRAPA-ultrasound imaging combination to detect vulnerable carotid plaque.

**Graphical Abstract:** 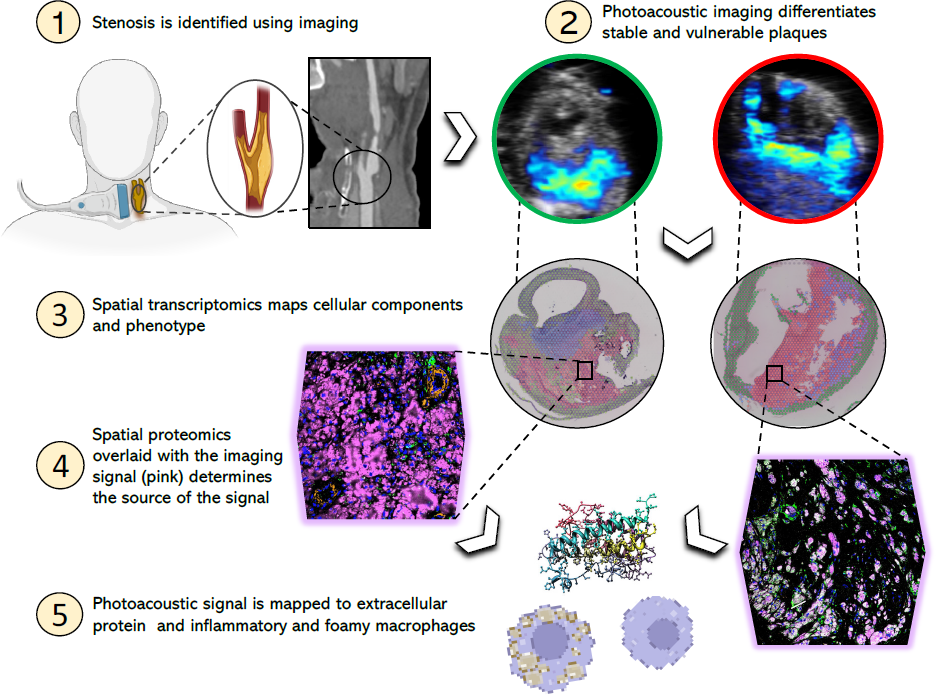

## INTRODUCTION

Cardiovascular diseases, which include coronary heart disease, peripheral arterial disease and stroke, are the leading cause of death globally ^1, 2^. In the United States, heart disease was the first and stroke the fifth leading cause of death in 2020^3^. The cause of these diseases in most cases is atherosclerosis^1^. Atherosclerosis is an inflammatory process resulting in the deposition of fatty and/or necrotic residues in the vessel wall and consequently the narrowing of the vessel lumen. The rupture of an atherosclerotic plaque and the following formation of a thrombus in the blood circulation can result in ischemic events such as myocardial infarction or stroke^1, 4, 5^. Silent, asymptomatic atherosclerosis is a common finding in the general population, even in young adults, and is typically associated with a low risk of myocardial infarction or stroke^6–8^; however, more than half of the acute coronary syndrome cases originate from these clinically silent plaques ^9–11^. For that reason, it is critical to understand the phenotypic characteristics of plaques, which can help develop imaging solutions for early detection ^12, 13^.

For the characterization of plaque types, specific histomorphological markers have been identified, including percent luminal stenosis, fibrous cap thickness, macrophage area, necrotic core area and calcified plaque area^14, 15^. The fibrous cap of an atherosclerotic plaque is one of the best discriminators of stable versus unstable plaque type^5, 14, 16, 17^. Standard imaging techniques to detect carotid atherosclerotic plaques are Doppler ultrasound, magnetic resonance (MR) angiography and computed tomography (CT) angiography. The clinical treatment process, including decisions about when to offer prophylactic surgery, is primarily based on the extent of luminal stenosis^18^. Novel imaging methods, including high-resolution MRI, CT combined with positron emission tomography (PET) imaging, and photoacoustic imaging are being tested to better assess the vessel wall features of the plaque in order to determine the plaques vulnerability^18–20^. Photoacoustic imaging utilizes laser light in the near-infrared range (680−980 nm) to excite endogenous or exogenous chromophores in the tissue in order to generate an ultrasound wave that can be combined with regular ultrasound scans^21^. Of particular clinical interest are endogenous chromophores used to acquire molecular tissue information without the requirement for an exogenous contrast agent. Specific endogenous tissue components including oxyhemoglobin (HbO_2_), deoxyhemoglobin (Hb), H_2_O, melanin and lipids can be identified via photoacoustic imaging based on their characteristic absorption spectra^22, 23^. Previous attempts to detect vulnerable plaque features via photoacoustic imaging focused on lipids (950-1250 nm)^24–26^ and intraplaque hemorrhage (808 nm)^27, 28^. Recently, bilirubin and other heme degradation products and insoluble lipid in atherosclerotic plaques have been evaluated as to their autofluorescence properties in the near-infrared (680 nm) range^29^. The hemoglobin degradation products represent a pathological process indicative of intraplaque hemorrhage and therefore serve as an important biomarker for vulnerable plaque ^29–31^. However, near-infrared autofluorescence (NIRAF) in the 650-700 nm range suffers from limited penetration depth and spatial resolution; and therefore, photoacoustic techniques are attractive ^22, 32^.

In this study, we investigate whether plaque components associated with the NIRAF signal can be detected with a clinical photoacoustic device to reveal the characteristics of a vulnerable plaque, such as thickness of a fibrous cap, infiltrating macrophages and necrotic core size. Further, we set out to characterize the molecular characteristics of the components that were responsible for the signal. To accomplish this, spatially registered near-infrared auto-photoacoustic (NIRAPA) and fluorescence microscopy are combined with immunohistochemistry, spatial RNA sequencing and immunofluorescence imaging via co-detection by indexing (CODEX) ^33^. Single-cell RNA sequencing has previously been successfully applied in atherosclerosis to profile individual cells and has revealed valuable information regarding infiltrating immune cells ^34, 35^; herein, we use spatial transcriptomics to associate NIRAF and photoacoustic signal with the gene expression profile. Spatial transcriptomics was performed with a depth of 15000 ∼ 18000 genes at each location, thus enabling correlation of the NIRAF signal with specific transcriptomic profiles^36^. Protein immunofluorescence, with single-cell resolution and markers spanning the NIRAF signal, immune and epithelial markers, was then applied to visualize the NIRAF signal generated by individual immune cells and extracellular components. The combination of these techniques (Extended Fig. 1) provides multiple levels of insight as to: 1) the association of the tissue level photoacoustic signal with clinically-significant features, 2) the characterization of an inflammatory gene signature that is primarily associated with the signal, and 3) confirmation that the photoacoustic and NIRAF signals result from extracellular matrix components and CD74^+^ HLA-DR^+^ CD14^+^ macrophages.

**Figure 1.**
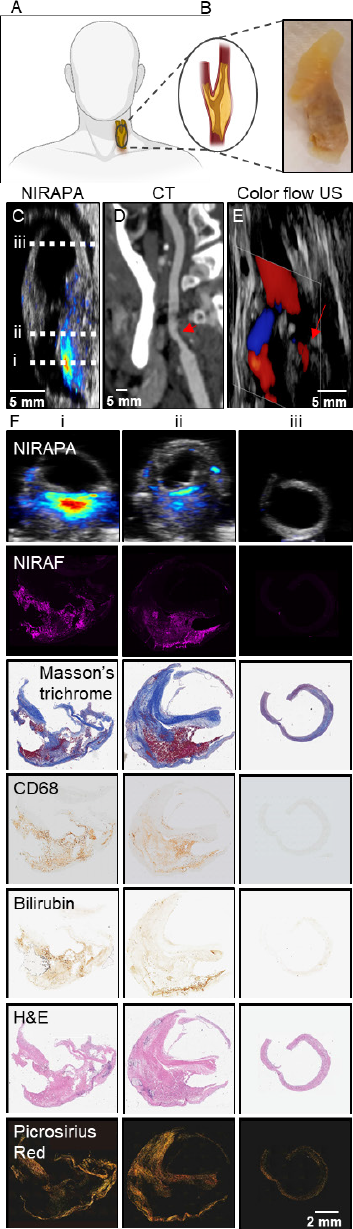
Photoacoustic imaging of the near-infrared auto-photoacoustic (NIRAPA) biomarker in human carotid plaque. A) Schematic of human carotid endarterectomy (CEA) sample. B) Human carotid plaque under white light. C-E) Longitudinal anatomic C) NIRAPA, D) computed tomography (CT) and E) color flow ultrasound (US) images of human carotid plaque. The dashed lines on the longitudinal image in (C) represent the imaging locations of the axial images in columns i, ii and iii in (F). F) Comparison of the NIRAPA signal (680-700nm) with NIRAF and Masson’s trichrome, CD68, bilirubin, H&E and picrosirius red staining for the three tissue section locations, i, ii and iii, indicated in Fig. 1C. Scale bar, bottom right, applies all panels in F.

## MATERIALS AND METHODS

### Human Carotid Endarterectomy Specimens

All human studies have been approved by the institutional review board at Stanford (IRB50541). To test the ability of spectroscopic photoacoustic (sPA) imaging to guide treatment decisions, human carotid plaques were used. Human carotid plaques were collected from 25 patients who presented to Stanford Hospital, Palo Alto, US with clinical indications for Carotid Endarterectomy (CEA), where 24 samples included carotid pathology and one sample was normal. In a subset of cases, contrast CT imaging and/or color flow imaging studies were available based on standard of care parameters and these studies were obtained and compared to the results. In each case, a region was scanned above and below the stenosis, such that each study contained a control region. Imaging was performed over a 15 to 39 mm distance at an inter-image distance of 0.2 mm, and the histology was acquired at an axial intersample distance of 1.5 cm. After surgical resection, the carotid specimens were immediately fixed in 4% paraformaldehyde (PFA) for subsequent photoacoustic imaging (PAI) and histopathologic assessment, or were imaged with PAI and fixed afterwards in 4% PFA. To evaluate the potential influence of PFA fixation on the PAI signal, these specimens were imaged again after fixation. A preliminary study confirmed that the NIRAPA signal was not impacted by fixation. sPA images and near-infrared auto-fluorescence (NIRAF) images were analyzed and compared with the results of the histopathologic analysis.

### Auto-near infrared photoacoustic imaging

Spectral photoacoustic images of the human carotid plaque were acquired using the Vevo LAZR-X (FUJIFILM VisualSonics) with a 15 MHz linear array transducer (MX 201; axial resolution, 100 um) and an average 150 mJ/cm^2^ average fluence laser pulse (10 ns pulse width, 20 Hz pulse repetition frequency, which has been optimized and calibrated). Single-plane, multiwavelength (680, 690, 700, 710, 720, 750, 800, 850 and 900 nm) photoacoustic images (Supplementary Fig.1) were acquired every 200 µm based on translation of a 3D stepper motor. B-mode ultrasound images were recorded simultaneously to provide anatomic registration. An example of the NIRAPA images acquired to generate the spectrum (Supplementary Fig. 1A) is provided as well as the estimated spectrum (labeled NIR-auto) for the plaque region (Supplementary Fig. 1B). The spectral amplitude of the NIRAPA signal (Supplementary Fig. 1B) is greatest in the lower NIR range (680-700 nm) and linearly decreases towards 950 nm. Spectral unmixing of the signal was computed using the oxygenated (OXY) and deoxygenated (DeOXY) hemoglobin settings on the Visualsonics yielding the estimated spectra and images (Supplementary Fig. 1B-D). Based on previous work demonstrating NIRAF from bilirubin ^29^, the bilirubin (B4126, Sigma-Aldrich) absorption spectrum was first measured using a VisualSonics phantom, where the intensity decreases with increasing wavelength and increases with bilirubin concentration (Supplementary Fig. 1C).

### Matching photoacoustic imaging (PAI) images to histologic sections

Samples were with the Vevo LAZR-X (FUJIFILM VisualSonics) for NIRAPA signal. To evaluate the clinical significance of the acquired PAI images, the corresponding histologic sections were examined in a blinded manner by a board-certified pathologist, and the images were examined by two experts in the field. The absence or thickness of the fibrous cap was recorded. Plaques with a fibrous plaque <65 µm or missing were classified as vulnerable plaques and plaques with a fibrous plaque >65 µm were determined as stable plaques^37^. The PAI reviewer assessed the fibrous cap using PAI images overlaid on top of B-mode ultrasound images. In each image, the NIRAPA signal was considered to be positive if the area was greater than 2.5 mm^2^. All cases included both positive images in the plaque center and negative imaging at the axial extrema of the excised vessel. A positive image separated from the vessel lumen by a hyperechoic ultrasound structure thicker than 65 µm was termed as stable plaque. A positive image adjacent to the vessel lumen with a smaller fibrous cap was assessed as vulnerable plaque. Afterwards, the results and clinical symptoms were matched. For each patient, a proximal and distal vascular region was identified and imaged to serve as an in-patient control. For the analysis of the PAI, all images were reviewed for each case and the minimum fibrous cap distance was quantified along with the maximum plaque volume.

### Spatial transcriptomic data processing

Five-µm sections from tissue blocks were placed on the Visium slides and subjected to spatial analysis using 10x Visium FFPE workflow (spatial resolution of 100 µm and with 1-10 cells per spot) (10x Genomics, Pleasanton, CA). Manufacturer’s instructions were followed without any significant alterations. Individually indexed libraries were pooled and sequenced on NovaSeq 6000 (Illumina inc., San Diego, CA) with the recommended read depth of per cell. Raw sequencing data was parsed through SpaceRanger analysis platform (10x Genomics), aligned with human (GRCh38) reference and low unique molecular identifier (UMI) counts were filtered. Transcriptomic analysis was performed with a Seurat framework ^38^. Separate samples were merged and then normalized with the SCTransform function, resulting in 3000-5000 spots for downstream processing. Principal component analysis was performed and, based on the elbow plot, the first 75 principal components were selected for downstream analysis. A resolution value of 0.4, k-nearest neighbors of 20, and the cutoff for Jaccard index of 0.005 were selected for Uniform Manifold Approximation and Projection (UMAP) clustering using the Leiden algorithm. To investigate cluster cell type, spatially variable features and cluster distinctive markers were determined with the FindSpatiallyVariableFeatures and FindAllMarkers function in the Seurat package^38^. The top differentially expressed genes of each clusters were compared to key gene markers from the literature to annotate clusters. The top differentially expressed genes from each sub-cluster were found with the FindAllMarkers function. All transcriptomic data processing, analysis and visualization was done with R language (version 4.2.2) in RStudio (RStudio team, PBC, Boston).

### CODEX processing

Plaque histology formalin-fixed paraffin embedded (FFPE) slices were stained with 51-multiplexed antibodies (Supplementary Table 4) and 3 spectral channels were co-acquired. After imaging processing, cell segmentation was performed with the DeepCell algorithm through the EnableMedicine portal (https://app.enablemedicine.com/portal). We further filtered the segmentation results based on size of the cell, total biomarker intensity, DNA channel intensity and signal coefficient of variation. The filtered segmentation results were then normalized and scaled for principal component analysis. UMAP clustering based on the Leiden algorithm was performed on 25 principal components with the number of k nearest neighbors, spread and minimum distance of clusters optimized to create a minimal number of clusters. To explore co-expressions of key immune markers (CD163, CD68, CD14, HLA-DR), we subset segmented cells with a normalized expression of larger than 2 and plotted a Venn diagram to visualize the population distribution of cells with various co-expression combinations. To explore co-expression signal level of key immune cell markers and NIRAF signal intensity, we computed the Pearson’s coefficient based on image signal intensity across the plaque for NIRAF signal intensity and the fluorescence signal intensity of immune cell markers such as CD14, HLA-DR, CD163, and Collagen IV as a negative control. Segmented cells were also exported to the cloud-based cytometry platform OMIQ (https://www.omiq.ai/) for additional visualization of marker fluorescence signal intensity.

### Statistics

Pearson correlation coefficients (r values) with estimated standard errors were used to determine associations between NIRAF signal and histologic measurements of CD68 and bilirubin. ImageJ – JACoP were used to calculate the Pearson correlation. An unpaired t test was used to evaluate the difference between the unstable plaque area NIRAPA signal and the stable plaque area NIRAPA signal. Simple linear regression was performed, and all graphs were created using GraphPad Prism version 9.3.1 for Windows (GraphPad Software, San Diego, California USA, www.graphpad.com). Sensitivity, specificity, positive predictive value and negative predictive value of PAI were determined with histologic analysis used as the reference standard.

## RESULTS

### NIRAF / NIRAPA signal correlates with macrophages and bilirubin

To assess the feasibility of using spectroscopic photoacoustic (sPA) imaging for the detection of vulnerable atherosclerotic plaques, 24 diseased human carotid plaques and one pathologically normal artery were collected from patients who underwent carotid endarterectomy (CEA) at Stanford Hospital, Palo Alto, USA. Twelve carotid plaques were collected from patients presenting with symptoms such as stroke or transient ischemic attacks, and 12 CEA samples were classified as asymptomatic (Fig. 1). The excised carotid (Fig. 1A-B) was subjected to NIRAPA imaging (Fig. 1C), where the images were oriented using CT (Fig. 1D) and color flow ultrasound (Fig. 1E) imaging. Most importantly, the NIRAPA images provide a positive contrast image of the plaque, as compared with the negative contrast produced by the absence of blood flow in CT and color flow ultrasound imaging. In all 24 diseased cases, the NIRAPA signal was detected in all patients in multiple imaging planes with the imaged region of interest spanning 15 to 40 mm (Fig. 1F).

A further example of the correspondence of the NIRAPA imaging and *in vivo* CT imaging, each acquired along the longitudinal axis, is provided in Supplementary Fig. 2. This clinical CT scan cannot distinguish between stable and unstable plaque components, whereas the photoacoustic scan can detect the NIRAPA signal and discriminate these different features. As a result, photoacoustic scanning is valuable for imaging the location of the stenosis and has clinical advantages when combined with other imaging modalities.

The acquired PAI cross-sectional images and corresponding histological sections and stains, spanning H&E, Masson’s trichrome, picrosirus red, CD68, and bilirubin (Fig. 1F), were registered to the acquired NIRAF image from the same slide, with the NIRAF signal representing the NIRAPA signal. A clear spatial correlation was observed between the NIRAPA and NIRAF signals, CD68, and bilirubin (Fig. 1F); therefore, we sought to quantify the correlation across the entire population in additional studies. Masson’s trichrome further defined regions of connective tissue and picrosirius red defined the collagen-rich regions.

### NIRAPA signal distinguishes between stable and unstable plaque

In order to evaluate the accuracy of photoacoustic images regarding discriminating stable plaque regions (a fibrous cap rich in collagen and alpha smooth muscle actin) versus unstable (vulnerable) plaque regions, the NIRAPA signal in the 680-700 nm range was compared to corresponding picrosirius red histological sections, indicating collagen-rich regions (Fig. 2). To evaluate the fibrous cap thickness, the distance between vessel lumen and unstable plaque region was measured using a cross-sectional ultrasound image overlaid with the NIRAPA signal of the carotid plaque. In the picrosirius red stain, the respective fibrous cap thickness was determined by measuring the distance between vessel lumen and signal-free areas within the vessel wall. If the fibrous cap was absent or undetected, we measured adjacent connective tissue areas and compared them to picrosirius red stain measurements. The measured fibrous cap thickness within the ultrasound-NIRAPA images correlated with the measured values from the picrosirius red histological images (r^2^ = 0.9913) (Fig. 2A). The same cross-section images were then used to evaluate the necrotic core area and the NIRAPA signal intensity difference between stable and unstable regions. The measured necrotic core areas based on NIRAPA signal corresponded to the signal-free picrosirius red stain areas (r^2^ = 0.8612) (Fig. 2B). Accordingly, the NIRAPA signal intensity recorded in unstable plaque areas was significantly higher than in stable plaque areas (p<0.0001, Fig. 2C).

**Figure 2.**
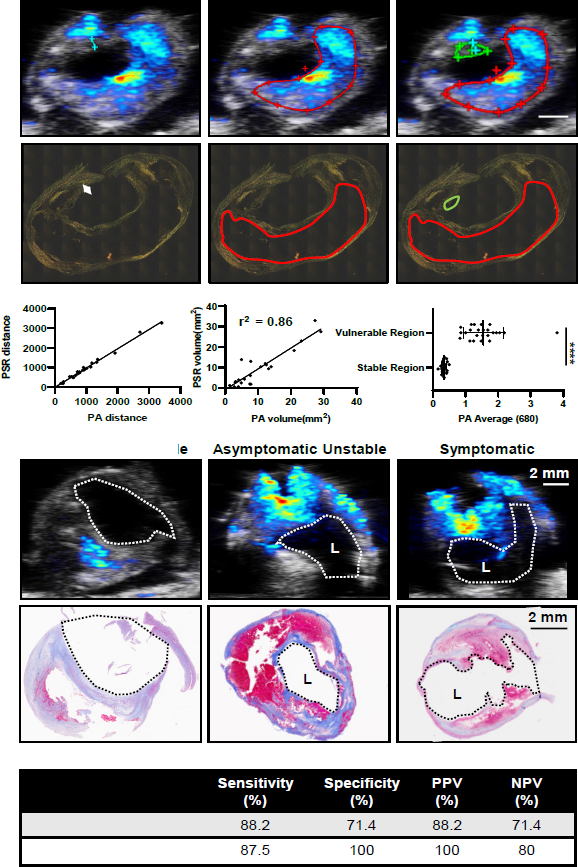
Measurements of fibrous cap thickness and plaque volume by histology and imaging establish sensitivity and specificity of the imaging technique. A) Measurements and correlation of fibrous cap thickness on NIRAPA (680-700 nm)-US images and picrosirius red (PSR) histopathology with r^2^=0.99. B) Measurements and correlation of the vulnerable plaque volume (red outline) on NIRAPA (680-700 nm)-US images and picrosirius red histopathology with r^2^=0.86. C) Photoacoustic (PA) signal intensity of NIRAPA (680-700 nm) averaged in vulnerable (red outline) and stable (green outline) plaque areas. Picrosirius red histopathology shows correlating areas. p<0.0001. D) Representative PA NIRAPA-US images of asymptomatic stable, asymptomatic vulnerable (unstable) and symptomatic vulnerable plaque cases. Correlating Masson’s trichrome stains. E) Summary of diagnostic accuracy of fibrous cap thickness as measured by photoacoustic imaging compared with histological classification. Vulnerable plaque defined as fibrous cap not existing or <65 µm. PPV: positive predictive value, NPV: negative predictive value, L: lumen, ****, p<0.0001. n=25 patients.

### NIRAPA plus ultrasound: sensitivity and specificity

The histological sections and NIRAPA-ultrasound images were then classified as stable or unstable/vulnerable, based on a fibrous cap thickness greater or less than 65 µm, respectively (Fig. 2D)^39^. In this comparison, NIRAPA images (using histology as a gold standard) achieved 88.2% sensitivity and 71.4% specificity (88.2% positive predictive value, 71.4% negative predictive value). In order to evaluate the clinical importance of NIRAPA images, we looked at asymptomatic cases separately. In 12 asymptomatic plaques, photoacoustic imaging reached 87.5% sensitivity and 100% specificity (100% positive predictive value, 80% negative predictive value) (Fig. 2E).

### Correlation of the NIRAPA signal with macrophages and bilirubin

We then sought to determine the origin of the NIRAPA signal. The naturally-occurring near-infrared signal correlated with the CD68 macrophage marker (Pearson’s coefficient 0.4, p<0.0001) and bilirubin (Pearson’s coefficient 0.4, p<0.0001), a degradation product of hemoglobin, in comparison with alpha smooth muscle actin (αSMA), a marker for a healthy and stable artery (Fig. 3A-B). Correspondingly, in the absence of a NIRAF signal, the detected area of CD68 and bilirubin (p<0.0001) was negligible and significantly lower than the αSMA area (Fig. 3A, C). This correlation with both macrophage localization and bilirubin concentration motivated us to further evaluate the molecular basis of the signal using spatial transcriptomics and proteomics.

**Figure 3.**
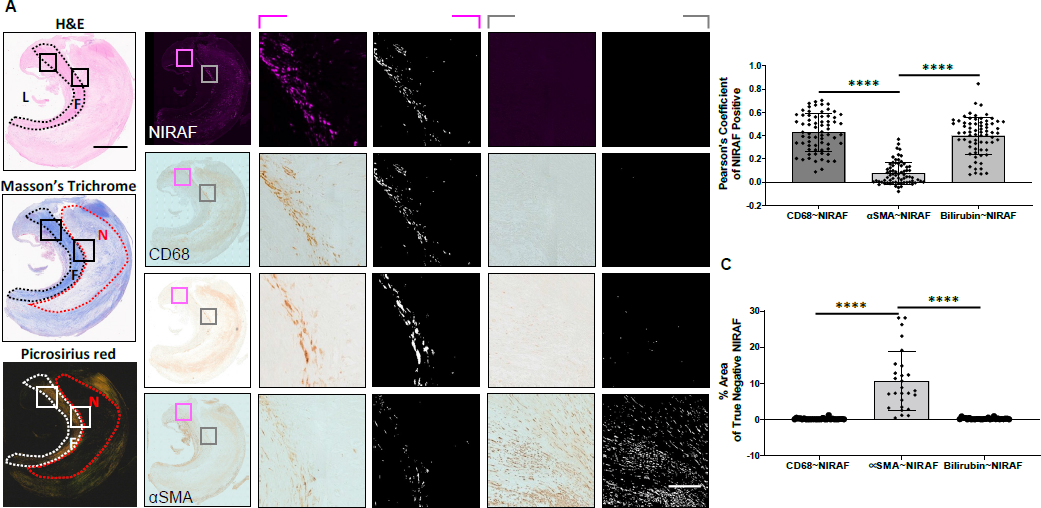
NIRAF signal correlates with CD68 and bilirubin. A) Overview of representative H&E, Masson’s trichrome and picrosirius red images used to localize the image features. CD68, bilirubin and aSMA images provided for reference. If NIRAF was detected, results were used to calculate the Pearson’s Coefficient (termed NIRAF True Positive, pink highlighted column). L: Lumen, F: Fibrous Cap, Necrotic: If NIRAF was not detected, the percentage pixel area was calculated (termed NIRAF True Negative, gray highlighted column). B) Pearson’s Coefficient results based on the colocalization of CD68, bilirubin and αSMA with respect to the NIRAF image. C) Difference in the analyzed pixel area of CD68, bilirubin or αSMA, when NIRAF was not detected. ****, p<0.0001, n=25 patients.

### Spatial transcriptomics and proteomic mapping in stable plaque

Imaging, immunohistochemistry (IHC), and spatial transcriptomic sequencing were performed on the plaque slide sections to probe the genetic character with respect to spatial location. As noted earlier, NIRAPA imaging corresponded closely with CT imaging and the location of the lesion (Supplementary Fig. 2). Based on the IHC analysis, we confirmed that the NIRAPA signal corresponded with both CD68 and bilirubin (Fig. 4A-D), where the bilirubin was localized to an area more than 1 mm from the lumen, and CD68 was detected within this region and a large surrounding area.

**Figure 4.**
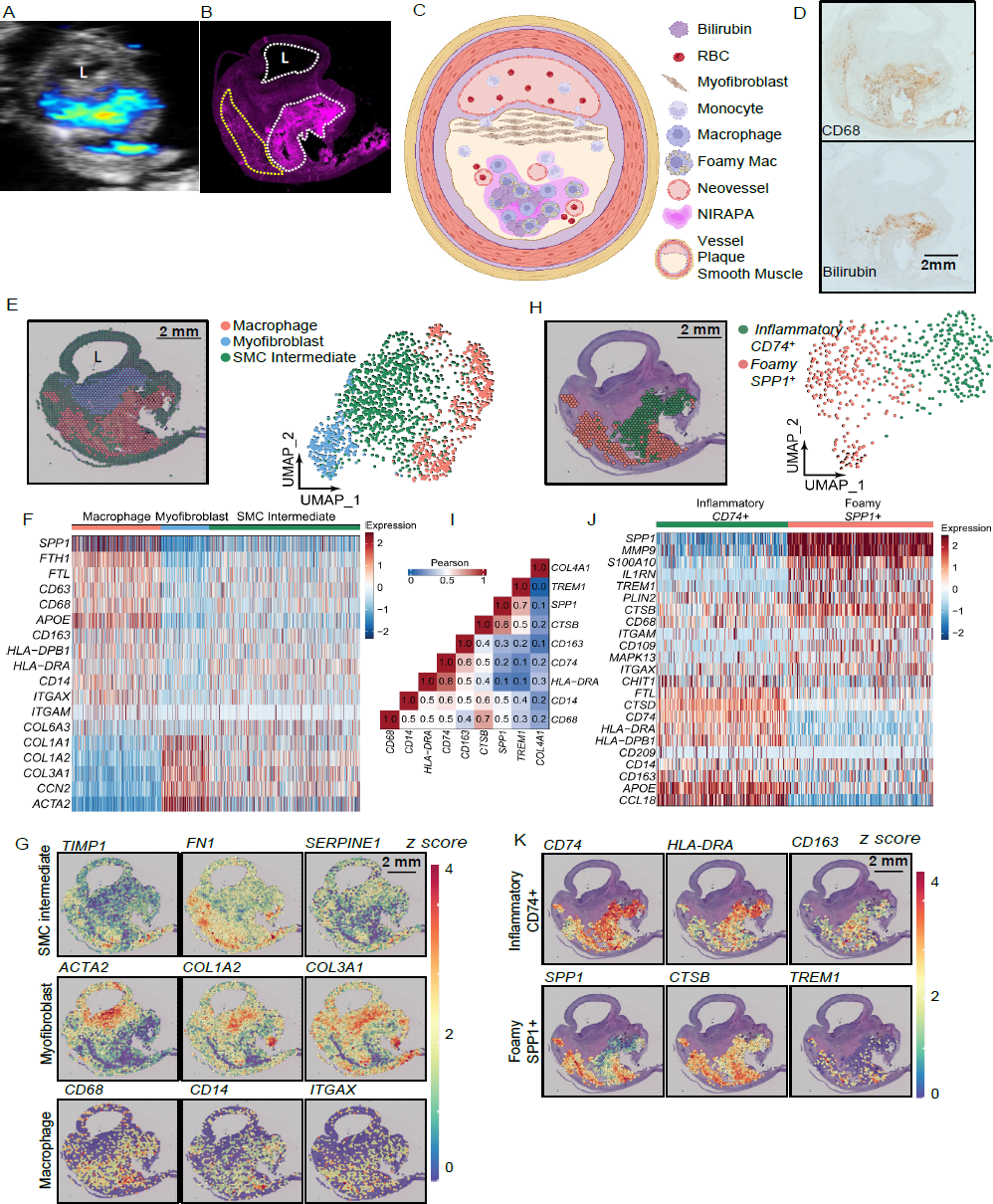
Spatial transcriptomic analysis of stable plaque specimen identifies specific macrophage populations that spatially correlate with the NIRAF and NIRAPA signals. A-B) NIRAPA (A) and NIRAF (B) images of a carotid plaque cross section. Annotations indicate strong (white) and weaker (yellow) NIRAF signal. C) Cartoon summarizing the stable plaque features and the location of the NIRAPA signal. D) Histological sections of the carotid endarterectomy (CEA) plaque specimen stained with CD68 and bilirubin. E) Overlay and Uniform Manifold Approximation and Projection (UMAP) cluster projection of spatial transcriptomics on carotid plaque H&E. Based on their gene expression, clusters have been assigned to macrophage, myofibroblast and smooth muscle cell (SMC) intermediate cell types. F) Overall heatmap of the general immunological signatures that differentiate the macrophage, myofibroblast and SMC intermediate clusters. G) Key genes that differentiate macrophage, myofibroblast, and SMC intermediate populations and their spatial intensity on the CEA specimen. H) Spatial deconvolution and UMAP cluster projection of the macrophage cluster in *CD74*^+^ and *SPP1*^+^ regions and the spatial location on the H&E-stained plaque cross section. UMAP projection of macrophage high resolution subtype clustering shows *CD74*^+^ and *SPP1*^+^ populations. I) Pearson’s correlation between genes within the macrophage clusters. J) Heatmap of macrophage-specific gene signatures that differentiate the *CD74*^+^ and *SPP1*^+^ macrophage subpopulations. K) Key genes differentiating inflammatory (*CD74*^+^) and foamy (*SPP1*^+^) macrophages and their spatial location on the CEA specimen. p value cutoff is 0.005. L: Lumen. Log_2_FC cutoff is 2.

With the Seurat single cell sequencing analysis package ^38^, we found 3 distinct clusters in the stable plaque. Through comparison between clusters and canonical cell markers commonly found in atherosclerotic plaque ^40–42^, the identified clusters included a smooth muscle-like phenotype (actin and collagen markers) surrounding the lumen, a myofibroblast cluster^41^ (*ACTA2*, *COL1A1*, *COL1A2*, *COL3A1*, and *CNN2*) in the proximal plaque and macrophages (*CD163*, *CD68*, *CD14*, *HLA-DRB1*, and *APOE*) in the distant plaque (Fig. 4E-F, Supplementary Table 1). Other groups have demonstrated that during atherosclerosis, smooth muscle cells (SMCs) transdifferentiate into fibroblast-like or macrophage-like cells^43^ and can undergo clonal expansion^44^. During transdifferentiation, SMCs begin to down-regulate SMC specific phenotypes. Since our first cluster expressed both SMC and macrophage phenotypes, but with a decreased expression, we annotated the first cluster as a SMC-derived intermediate. Comparing the spatial transcriptomic results (Fig. 4E) with NIRAPA and NIRAF data (Fig. 4A-B)), a segment of the macrophage population overlapped with the intense NIRAF signal, and a population of macrophages exhibited with a greatly reduced NIRAF signal. Maps of the spatial distribution of key markers (Fig. 4G) further defined the spatial characteristics.

Since sequencing and NIRAF data indicated potential sub-macrophage populations, we computationally isolated the macrophage cluster and further re-clustered with a higher resolution, following Seurat’s standard clustering protocol (Fig. 4H). Through high-resolution re-clustering, we discovered two distinct macrophage subpopulations, which differentially expressed key gene markers (*CD74* and *SPP1*) (Fig. 4H). We then quantified the spatial correlation between the expression of various macrophage markers (Fig. 4I), the correlation between inflammatory markers was greatest (0.8) between *HLA-DRA* and *CD74* and was similarly large within the *SPP1* cluster. Based on select differentially-expressed genes of each cluster, we found that macrophages with a greater imaging signal and higher *CD74* expression also co-expressed MHC II (*HLA-DRA*) and *APOE*, while macrophages with higher *SPP1* expression co-expressed *S100A10*, *MMP9*, *CTSB*, *IL1RN*, and *TREM1* (Fig. 4J). The cluster co-expressing macrophage activation markers such as *CD74*, *APOE*, and *HLA-DRA* spatially overlapped with the greater NIRAPA and NIRAF signals (Fig. 4K) with a significant correlation between the NIRAF signal level and that of *CD74*, *CD163*, and *HLA-DRA* (Supplementary Fig. 3) ^45^. In contrast, the macrophage cluster co-expressing *SPP1* and *CTSB* spatially overlapped with the reduced NIRAF signal (Fig. 4K, Supplementary Fig. 3). *CTSB* is a cathepsin known to promote atherosclerotic inflammation and vulnerability^41^ (Fig. 4K) and the *SPP1* and *CTSB* cluster is associated with foamy macrophages with an M2-like phenotype^41^.

Since spatial transcriptomics identified inflammatory macrophage populations related to the NIRAF signal, we probed the spatial protein distribution and the relationship between protein expression and the NIRAF signal at the single-cell level in stable plaque (Fig. 5). We performed spatial proteomic imaging on slices adjacent to the spatial transcriptomic studies and acquired the coincident NIRAF signal. Spatial correlation between the NIRAF signal and MHCII (HLA-DR) and CD14 was 0.9 and 0.8, respectively (Fig. 5A-C), and HLA-DR and CD14 spatially correlate with one another at 0.9. As expected, the correlation of the NIRAF signal with Collagen IV was low (0.2), and in this stable plaque, the correlation with CD163 was 0.2 (Fig. 5D). We segmented the cells based on the DAPI nuclear DNA stain and plotted cell populations expressing each marker in a Venn diagram (Fig. 5E). The greatest marker overlap occurred between CD14 and HLA-DR, with ∼20% of the segmented cells displaying both markers.

**Figure 5.**
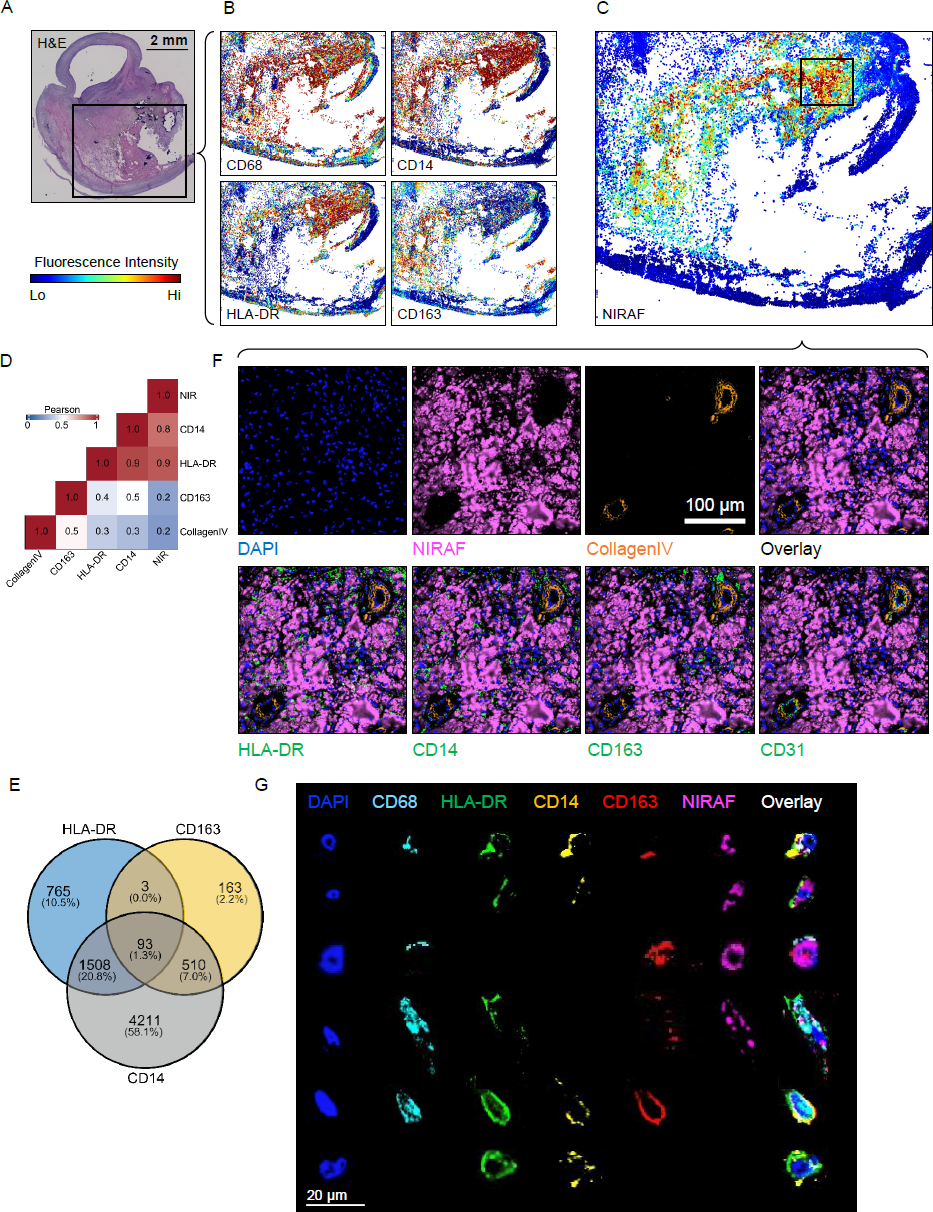
Spatial proteomic analysis of resolved single cells and correlation with overlaid NIRAF signal in a stable plaque. A) H&E cross section overview with black box showing CODEX region. B) CODEX images showing signal intensities of CD68, CD14, HLA-DR and CD163 on the same tissue section. C) CODEX image of NIRAF signal intensity on the same tissue section with the high-NIRAF signal region highlighted and the boxed region studied further in F. D) Pearson’s correlation between the NIRAF signal, genes within the macrophage clusters and Collagen IV. E) Venn diagram of the expression of HLA-DR, CD163 and CD14 based on segmented cells from CODEX images. F) ROI images of CODEX showing DAPI, NIRAF, Collagen IV, HLA-DR, CD14, CD163 and CD31. G) Representative manually-segmented individual cell fluorescence examples from CODEX.

To further characterize the source of the NIRAF and NIRAPA signal, we examined the CODEX images with successively higher spatial resolution (Fig. 5F-G). As expected from Fig. 4A-D (bilirubin distribution), we found that in some regions the NIRAF signal correlated with extracellular protein (regions of pink fluorescence in the absence of a DAPI signal). Individual neovessels were identified deep within the plaque, particularly in the NIRAPA region. The NIRAF/NIRAPA signal was frequently associated with the presence of CD31^+^ angiogenic neovessels, which can facilitate red blood cell extravasation (Fig. 5F, Collagen IV, CD31).

Signal overlap with the HLA-DR and CD14 markers was also detected and then probed at higher spatial resolution in Fig. 5G. Individual cells were manually segmented and images were obtained with a set of markers. The analysis confirmed that the signal was localized with the cytoplasm of CD68^+^ macrophages with varied expression of HLA-DR, CD14, and CD163. This analysis also demonstrated that the markers could be detected in these same regions without the presence of the NIRAF signal, suggesting that the varied cellular contents and/or phagocytotic activity determine the strength of the NIRAF signal.

We next evaluated the spatial correspondence of the transcriptomic and proteomic signals (Supplementary Fig. 4) in the structural features of the plaque. We found good agreement between the key protein/gene pairs of interest; the αSMA/*ACTA2* signal was confirmed and correlated, the CD31/*PECAM1* signal was correlated and the Collagen IV/*COL4A1* signal (which largely corresponds with the blood vessels) was mapped. This is a particular advantage of combining these techniques, in which spatial proteomics provides single-cell spatial resolution to detect features such as angiogenic vessels, and spatial transcriptomics provides high-depth profiling of global gene expression.

In summary, the CODEX analysis of the stable plaque cross section confirmed the spatial RNA sequencing results on a single-cell level. The NIRAF signal was spatially colocalized in some pixels containing extracellular matrix without DAPI (spatially correlated with bilirubin) and individual inflammatory cells expressing HLA-DR, CD14 and CD163.

### Spatial transcriptomic sequencing and proteomics highlights inflammatory macrophages lining the lumen of vulnerable plaque

We then evaluated the source of the signal in a vulnerable plaque (Fig. 6) where the corresponding CT and NIRAPA images are summarized in Supplementary Fig. 5 and the associated videos. Here again, the NIRAPA and NIRAF signals were similar and spatially localized within this vulnerable plaque. The NIRAPA signal extended a smaller distance from the lumen, possibly due to the presence of proximal calcification blocking light from entering the intact plaque. From Fig. 6A, the combination of the ultrasound and NIRAPA signal suggested a very thin fibrous cap, and this was confirmed in Fig. 6B, as illustrated in Fig. 6C. Both CD68 and bilirubin IHC were positive for the region of the active signal (Fig. 6D-E). Cluster analysis of the spatial transcriptomic data again detected the presence of three clusters spanning macrophages, myofibroblasts and an SMC intermediate cluster (Fig. 6F-G, Supplementary Table 2). Here, the macrophage cluster was adjacent to the lumen (Fig. 6F). The macrophage cluster was further analyzed to reveal foamy, *APOE*^+^ and inflammatory subclusters (Fig. 6H), with a thin layer of inflammatory macrophages covering the lumen and correlated with the NIRAPA signal. This inflammatory cluster highly expressed *CD74*, *HLA-DRA*, *CD14*, and *CD163* (Fig. 6I). Spatial mapping of individual genes differentiated the macrophage clusters (Fig. 6J), with a region with enhanced *CD74* expression outlined. CODEX imaging focused on this region revealed expression of angiogenic and inflammatory macrophage markers (Fig. 6K).

**Figure 6.**
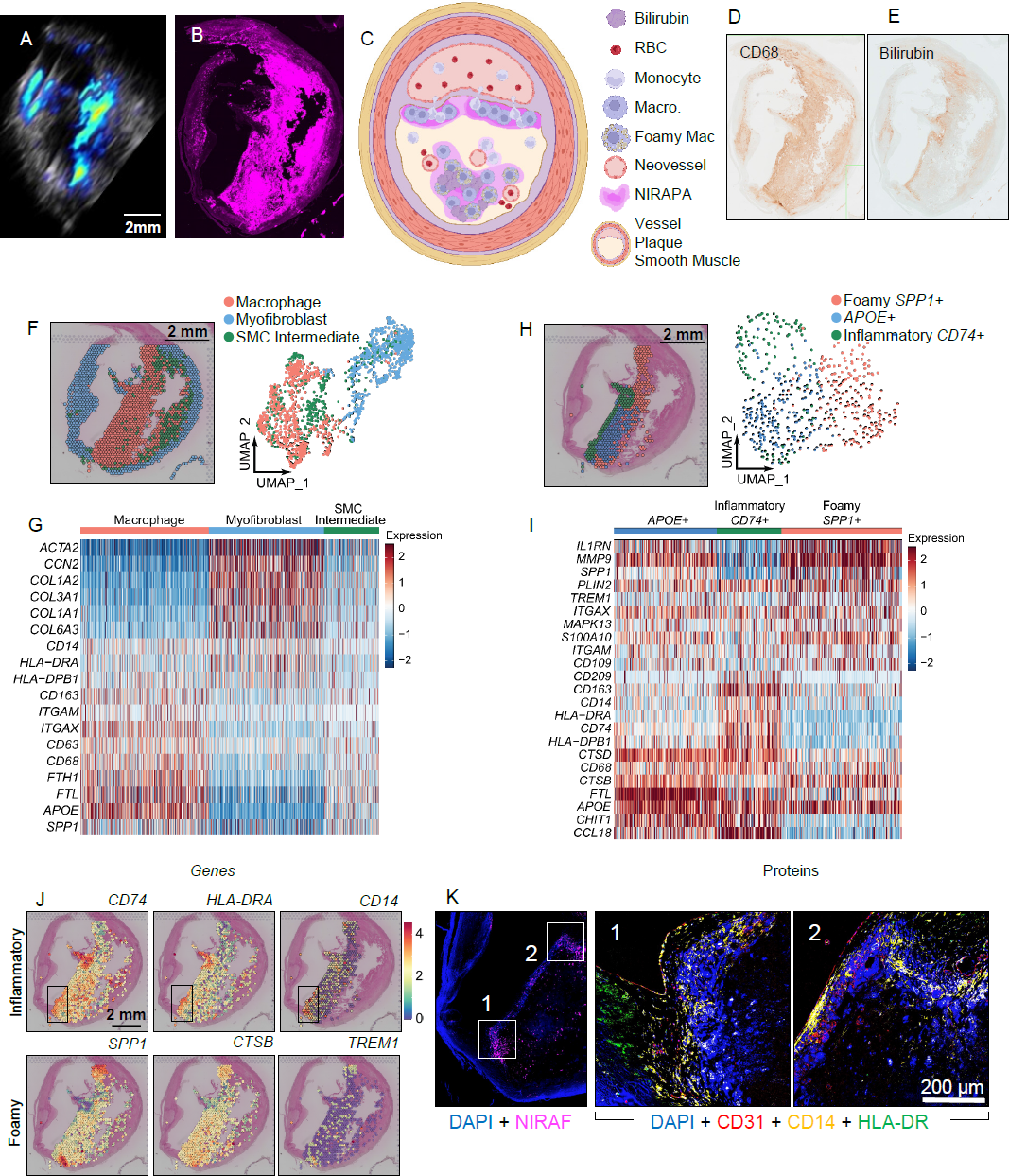
Spatial transcriptomic and proteomic analysis of vulnerable plaque delineates spatially-dependent macrophage populations. A-B) NIRAPA (A) and NIRAF (B) images of a carotid plaque cross section. C) Cartoon summarizing the vulnerable plaque features and the location of the NIRAPA signal. D-E) Histological sections of the carotid endarterectomy (CEA) plaque specimen stained with CD68 (D) and bilirubin (E). F) Overlay and Uniform Manifold Approximation and Projection (UMAP) cluster projection of spatial transcriptomics on carotid plaque H&E. Based on their gene expression, clusters have been assigned to macrophage, myofibroblast and smooth muscle cell (SMC) intermediate cell types. G) Overall heatmap of general immunological signatures that differentiate the macrophage, myofibroblast and SMC intermediate clusters. H) Spatial deconvolution and UMAP cluster projection of the macrophage cluster in inflammatory *CD74*^+^, foamy *SPP1*^+^, and *APOE*^+^ regions and the spatial location on the H&E-stained plaque cross section. UMAP projection of macrophage high resolution subtype clustering shows *CD74*^+^, *SPP1*^+^, and *APOE*^+^ populations. I) Heatmap of macrophage-specific gene signatures that differentiate the *CD74*^+^, *SPP1*^+^, and *APOE*^+^ macrophage subpopulations. J) Key genes differentiating inflammatory (*CD74*^+^) and foamy (*SPP1*^+^) macrophages and their spatial location on the CEA specimen. A region with enhanced *CD74* expression is outlined in a black box overlay and investigated further in (K). K) CODEX imaging of DAPI nuclear DNA stain, NIRAF, CD31, CD14, and HLA-DR in two areas of the vulnerable plaque region showing enhanced *CD74* expression. P value cutoff is 0.005. Log_2_FC cutoff is 2.

The analysis of this vulnerable plaque was repeated in a second slice (Supplementary Fig. 6, Supplementary Table 3) and confirmed the correspondence of the NIRAPA and NIRAF signals with a macrophage (CD68) and bilirubin signal (Supplementary Fig. 6A-E), the existence of the inflammatory macrophage mRNA near the lumen (Supplementary Fig. 6F-I). The co-expression of CD14 and HLA-DR on ∼20% of cells was observed (Supplementary Fig. 6J). Finally, in this vulnerable plaque, the expression of the NIRAF signal within the cytoplasm of individual macrophages was confirmed on CODEX imaging (Supplementary Fig. 6K). In summary, the NIRAPA technique was capable of detecting regions of vulnerable plaque as a result of signals generated by extracellular protein and macrophages.

## DISCUSSION

Standard imaging modalities for assessing carotid artery atherosclerosis include sonography, CT, MR angiography, and digital subtraction angiography^46^. Treatment decisions, including determining when to offer surgical intervention, are primarily based on the degree of stenosis and the presence or absence of clinical symptoms.^47^ However, plaque size and the severity of stenosis are not necessarily correlated with plaque vulnerability.^37^ Many recent studies emphasize the importance of the plaque composition, which has a significantly higher impact on plaque vulnerability than luminal stenosis and plaque size alone.^48–51^ Unstable, or vulnerable, plaques are characterized by a thin fibrous cap, a large necrotic core, neovascularization from vasa vasorum, and intraplaque hemorrhage ^15, 52–54^. CT is part of the current guidelines for the assessment and management of carotid plaques. The degree of stenosis in combination with clinical symptoms still guide the treatment decision. However, CT imaging alone cannot assess plaque composition ^55, 56^, nor reliably differentiate fibrous tissue and intraplaque hemorrhage due to the overlapping Hounsfield units of these components^57^.

High-resolution MRI is used to accurately detect intraplaque hemorrhage, a marker of plaque vulnerability in symptomatic and asymptomatic patients^51, 58^. However, MRI is the most expensive imaging technique and not broadly accessible, whereas photoacoustic imaging (PAI) in combination with ultrasound can be integrated more broadly ^21^. Further, mapping the fibrous cap thickness is challenging with MRI imaging. PAI, utilizing our proposed auto-photoacoustic signal, would allow physicians to assess plaque vulnerability quickly and affordably, thus potentially facilitating the identification of asymptomatic individuals requiring treatment. This could lead to a significant reduction of adverse outcomes, as the presence of intraplaque hemorrhage is an independent risk factor for stroke and coronary heart disease^51^.

For these reasons, new clinically-relevant imaging techniques such as PAI, in combination with novel biomarkers, are needed to further improve plaque treatment regimens. In our study, we use photoacoustic imaging to discriminate stable versus vulnerable plaque components based on naturally-occurring near-infrared markers. Near-infrared autofluorescence (NIRAF) has been associated with lipids and intraplaque hemorrhage. Macrophages, which are a marker of a vulnerable plaque, phagocytose extravasated red blood cells, degrade heme to bilirubin, and are involved in the formation of insoluble lipids or ceroids ^29, 31^. Here we show that the aforementioned endogenous near-infrared biomarkers, especially in colocalization with macrophages, can be detected by PAI. Our results demonstrate that the NIR-auto-photoacoustic (NIRAPA) signal in the 680-700 nm range can be combined with anatomic ultrasound to distinguish stable from vulnerable plaque components. Especially in asymptomatic cases where the need for surgery is still difficult to assess, PAI using the NIRAPA signal achieved significant sensitivity of 87.5% and specificity of 100% (n=12). The sensitivity and specificity achieved with PAI outperformed the classical symptoms/histology system. The higher sensitivity and specificity could also be achieved due to the identification of asymptomatic vulnerable plaques, which represent the most difficult to detect and potentially most clinically-relevant cases.

Of particular clinical relevance is the assessment of the fibrous cap thickness overlying the necrotic core, which is a key source of inflammation and thrombogenicity in lesions at risk for erosion or frank rupture^59^. A fibrous cap measuring less than 65 µm is considered a vulnerable plaque^37^. Photoacoustic images overlaid on B-mode ultrasound images, with a resolution of 100 µm, allowed us to identify thick (>65 µm) fibrous caps, based on the missing NIRAPA signal, and NIRAPA signal generating pathological tissue components (macrophages, hemoglobin degradation products, necrotic material). The clinical importance of the detected NIRAPA signal depends on its area and its localization with respect to the plaque lumen. Some MRI-based studies have indicated that debris with a volume larger than 100 mm^3^ can promote blood vessel occlusion and cause stroke^60^.

Spatial transcriptomics is innately quantitative and was applied to cluster and assess the smooth muscle and macrophage cell populations. With this technique, a layer of inflammatory macrophages and angiogenic vasculature was detected on the luminal surface of the vulnerable plaque. The spatial RNA sequencing results, in combination with spatial proteomics and IHC, confirmed spatial correlation of the NIRAF signal and macrophages as well as hemoglobin degradation products^29, 61^. Additionally, both techniques revealed that inflammatory macrophages (*CD74*, *HLA-DRA*) were associated with a stronger NIRAF signal than *SPP1*^+^ foamy macrophages.

By combining spatial transcriptomics and proteomics, we were also able to precisely determine the source of the NIRAPA signal. First, CODEX imaging with the NIRAF signal overlay confirmed the macrophage phenotypes associated with the NIRAPA signal on a single-cell level. Indeed, in a manner similar to imaging cytometry, the localization of the NIRAPA signal within the macrophage cytoplasm was confirmed. Second, the CODEX imaging mapped angiogenic vascular structures at a resolution not feasible with Visium transcriptomics, where leaky, angiogenic vessels could be the source of the red blood cells and their degradation products. While additional spatial transcriptomics technologies are emerging, few will be able to provide single-cell resolution with a deep genetic profile, further emphasizing the power of combinatorial-omics analyses. Third, with spatial proteomics, we confirmed that extracellular protein was a partial source of the NIRAPA signal.

To compensate for the patient’s differential light penetration properties, the PAI protocol should begin in a vessel area without plaque. In this area, PAI can be calibrated and the laser intensity or wavelength summation determined. Based on this PAI configuration, the operator should continue scanning the plaque area. PA images within the NIR range of 680-700 nm overlaid on top of B-mode ultrasound images then allow the assessment of the plaque wall. The size and location of the NIRAPA signal with respect to the plaque lumen can be used to distinguish the respective plaque as stable versus vulnerable. Differences between the NIRAPA signal and NIRAF are influenced by the plaque size and the resulting illumination limitations of the whole plaque sample. This physical limitation of the current photoacoustic devices may require the development of novel transducers, which circumvent the respective carotid artery and therefore allow more holistic illumination and detection of plaque components.

Atherosclerosis is a dynamic process over time, which is characterized through different stages. Since plaques are very common among aging adults and the majority consist of stable plaques which do not impact the wellbeing of a patient, it is essential to identify and prioritize treatment of vulnerable plaques. Carotid artery plaque management requires thorough surveillance due to the potential for devastating morbidity from stroke. Depending on the plaque characteristics, different treatment strategies are indicated, ranging from intensification of drug regimens to surgical interventions. To aid in predicting which treatment strategy will be the most successful, the detection of this newly described naturally-occurring near-infrared biomarker, which is associated with the presence of inflammatory markers such as macrophages and the blood degradation product bilirubin, is crucial. Therefore, our results will lead to more precision medicine and personally-tailored disease evaluation and treatment.

## Data Availability

All data produced in the present study are available upon reasonable request to the authors

## Acknowledgements

We dedicate this paper to our friend and colleague Dr. Sanjiv Sam Gambhir who recruited Martin Schneider to Stanford University. This project was supported by the Stanford PHIND Institute and NIH R01 CA250557 and NIHR01CA253316 through which we developed spatial sequencing techniques. The sequencing data was generated with instrumentation purchased with NIH funds: S10OD025212, 1S10OD021763, R35HL144475 and T32CA118681. This research was also supported in part by a training grant from NIH Cellular and Molecular Training Grant (NIGMS, T32GM007276). The Vevo LAZR-X equipment was provided for this study by Fujifilm VisualSonics. The assistance of Alexander Trevino at Enable Medicine was greatly appreciated.

## Competing interests

AM is an employee of EnableMedicine which develops CODEX-related methodologies. All other authors declare no competing interest.

## Data availability

The main data supporting the findings of this study are available within the paper and its Supplementary Information. The raw and analyzed datasets will be made available through an appropriate data server upon acceptance of the paper.

## Author contributions

M.K.S., J.W., N.J.L., J.J.H., K.W.F conceived and designed the experiments. M.K.S., J.W., D.W., J.C., S.S.A., C.F.B., T.A., G.K.S, S.J.S, A.M., performed the experiments. M.K.S., J.W., A.K., D.S., S.S.A., A.B., S.R.L, M.G.L, N.J.L, J.J.H., K.W.F. analyzed the results. M.K.S., J.W., A.K., K.W.F. wrote the manuscript. K.W.F supervised the entire project. All authors discussed the results and commented on the manuscript.

## Notes

### Competing Interest Statement

Aaron Mayer is an employee of EnableMedicine which develops CODEX-related methodologies. All other authors declare no competing interest.

### Author Declarations

All human studies have been approved by the institutional review board at Stanford (IRB50541).

