## Supplement for "Combined near infrared photoacoustic imaging and ultrasound detects vulnerable atherosclerotic plaque"

**for**

**List of contents:**

Supplementary Methods.

Supplementary Fig. 1. Spectrum of the NIRAPA signal and the bilirubin correlation.

Supplementary Fig. 2. Comparison of contrast CT images PAI images using the NIRAPA signal to detect vulnerable plaque composition of a stable carotid artery.

Supplementary Fig. 3. Quantification and statistical comparison of the key gene counts corresponding to NIRAF signal regions.

Supplementary Fig. 4. Spatial correspondence of the transcriptomic and proteomic signals

Supplementary Fig. 5. Comparison of contrast CT images with PAI images using NIRAPA signal to detect vulnerable plaque compositions of a vulnerable carotid artery.

Supplementary Fig. 6. Spatial transcriptomic and proteomic analysis of vulnerable plaque delineates spatially-dependent macrophage populations.

Supplementary Table 1. Differential expression gene and location coordinate tables for the CEA sample from Fig. 4. Including overall populations and macrophage subpopulations

Supplementary Table 2. Differential expression gene and location coordinate tables for the CEA sample from Fig. 6. Including overall populations and macrophage subpopulations

Supplementary Table 3. Differential expression gene and location coordinate tables for the CEA sample from Supplementary Fig. 6. Including overall populations and macrophage subpopulations

Supplementary Table 4. CODEX proteins

### Supplemental Methods

#### Auto-near infrared photoacoustic imaging

The lateral resolution of the Vevo LAZR-X imaging system with a 15-MHz (10–22 MHz) linear-array transducer was 100 µm, which also corresponds to the lateral spatial resolution of the transcriptomic data. In order to reduce ultrasound reflections, the human carotid plaque was placed on fixed gauze in a custom-made polyvinyl chloride plastisol (Item No. 8228SS, M-F Manufacturing Company) vial. 0.35% intralipid (CAS Number: 68890-65-3, Sigma-Aldrich) was dissolved in phosphate buffered saline (PBS) to simulate tissue-specific light scattering properties.

#### Immunohistochemistry

5 µm tissue sections were stained with hematoxylin and eosin (H&E), Masson's trichrome (Sigma Aldrich) and picrosirius red (24901; Polysciences, USA). Picrosirius red sections were imaged using a polarized light (AxioImager Widefield Fluorescence Microscope, Zeiss).

After the plaques have been fixed in 4% PFA and paraffin embedded, 5 µm slices were sectioned from the tissue block. Paraffin was removed using Xylene and then hydrated deploying an alcohol/water gradient. The de-paraffinized slides were air dried and imaged for the NIRAF signal using a digital slide scanner (CY5, Olympus VS120, exposure time 500 ms, 20x objective). The staining protocol was continued by antigen retrieval in citrate buffer (cat. no.: C9999, Sigma-Aldrich, USA) for 20 min at 95 degrees Celsius. After the slides reached room temperature, they were washed using a wash buffer (PBS + 0.1% Tween) and subsequently treated with 3% hydrogen peroxide for 10 min to block endogenous peroxidase activity. Following an additional washing step in wash buffer, the sections were incubated with goat serum (Vector Laboratories, CA, USA) for 30 min. The sections were then covered with a primary antibody and incubated overnight at 4 degrees Celsius. Anti-CD68 (clone KP1, BioLegend, USA, dilution 1:250), anti-alpha-SMA (cat no: ab5694, Abcam, USA, dilution 1:200) and anti-bilirubin (clone 24G7, Shino-test, Kanagawa, Japan, dilution 1:200) were used as primary antibodies. After washing off the primary antibody, the sections were incubated with a corresponding biotinylated secondary antibody for 30 min at room temperature. Following an additional washing step, the sections were incubated with a DAB (3, 3'-diaminobenzidine) substrate kit for peroxidase detection (Vector Laboratories). No counterstaining was performed to achieve maximal contrast for digital imaging processing.

#### Cluster annotation

The macrophage cluster was isolated and further sub-clustered with a resolution value of 0.8, 20 k-nearest neighbors, and a Jaccard index cutoff of 0.005. The macrophage cluster was annotated based on the expression of *CD68*, *CD14*, *HLA-DRB1*, *HLA-DRA*, and *APOE*. The *ACTA2*$^{+}$ myofibroblast cluster was based on the expression of *ACTA2*, *COL1A1*, *COL1A2*, and *CNN2*. Pro-inflammatory macrophages were defined by the higher expression of *CD74, HLA-DRA,* and *APOE* (*39*). Foamy macrophages were defined by the higher expression of *CTSB*, *TREM1*, *S100A10,* and *SPP1* (*39*). A cluster, with combined weak expression of *COL1A1*, *COL3A1*, *CD68*, *CD14* and *SPP1,* was categorized as smooth muscle cell (SMC) derived intermediate cell types (*38*).

### Supplementary Figures and Tables

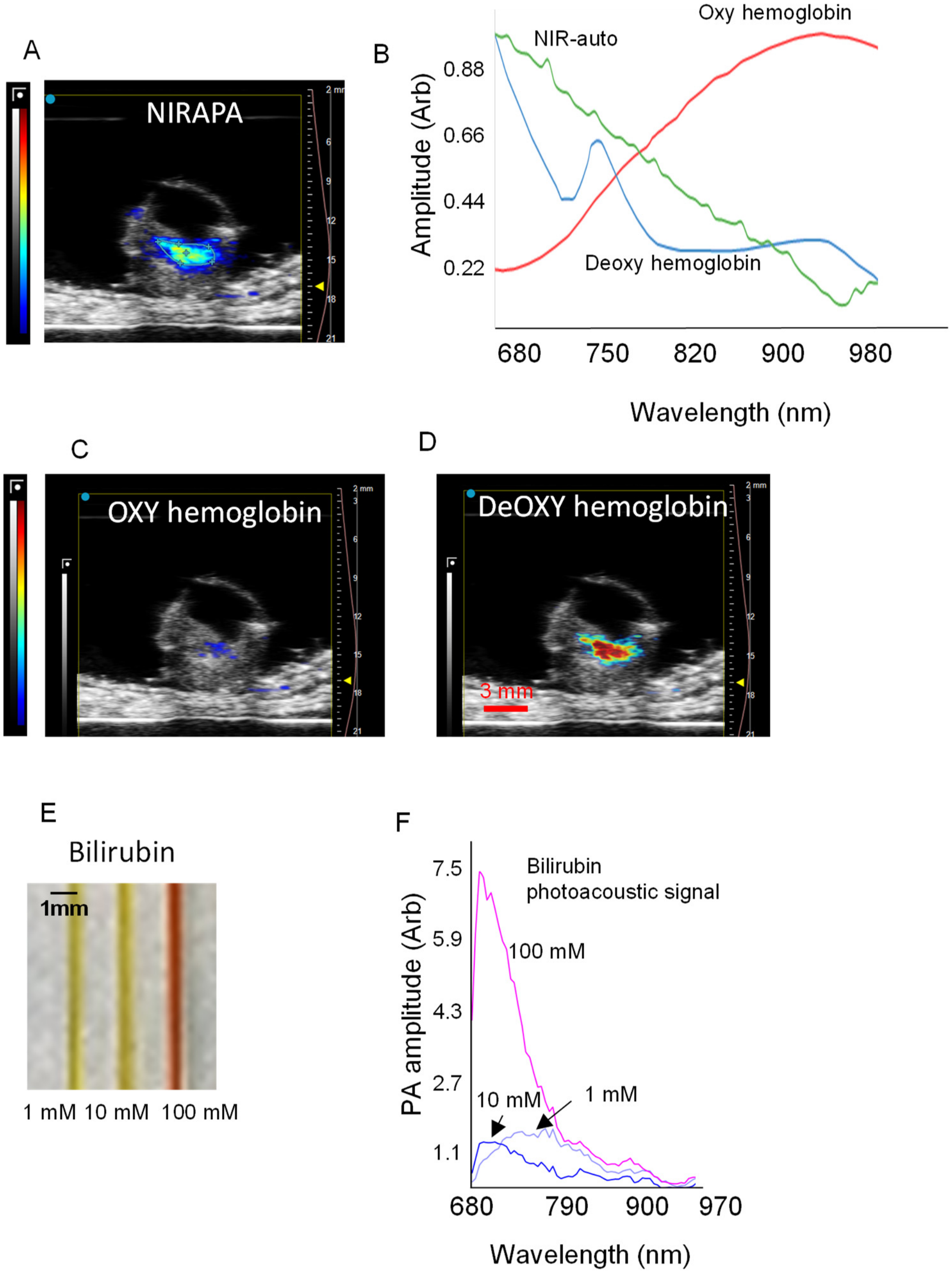


**Supplementary Fig. 1. Spectrum of the NIRAPA signal and the bilirubin correlation.** A) Near-infrared Auto-Photoacoustic (NIRAPA) sample image. B) Spectra of Near-infrared Auto-Photoacoustic (NIRAPA) and Vevo unmixing into spectra for oxyhemoglobin ($HbO_2$) and deoxyhemoglobin (Hb). C-D) Spectrally unmixed images of C) Oxyhemoglobin ($HbO_2$), D) Deoxyhemoglobin (Hb) calculated by Vivo. E) Photoacoustic phantom containing 1 mM, 10 mM and 100 mM of bilirubin in DMSO. F) Photoacoustic signal spectra of different bilirubin concentrations.

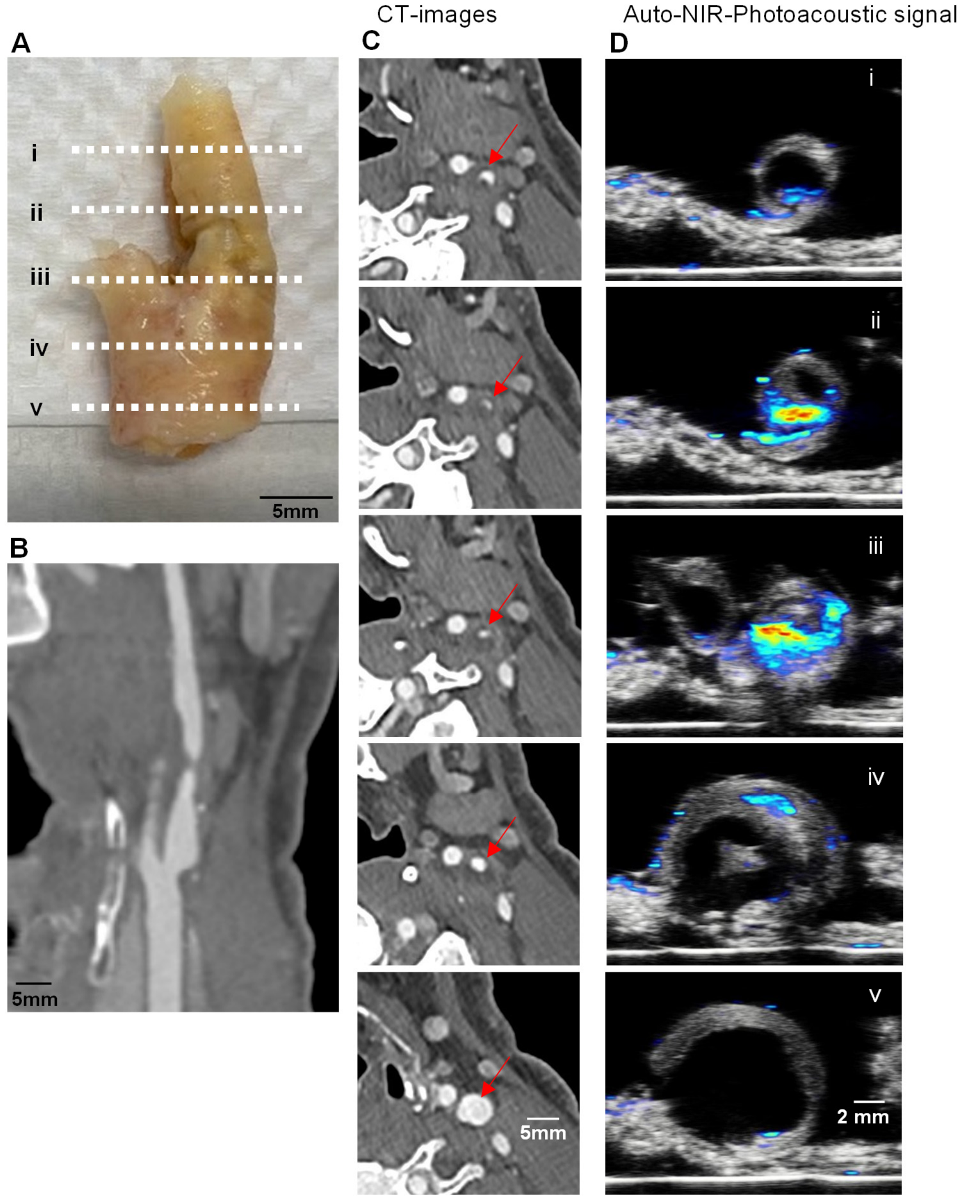


**Supplementary Fig. 2. Comparison of contrast CT images with PAI images using the NIRAPA signal to detect vulnerable plaque composition of a stable carotid artery.** A) Human carotid endarterectomy (CEA) specimen under white light. B) Corresponding longitudinal contrast CT image shown underneath. The dashed lines on the longitudinal image represent the imaging locations of the axial images in columns B and C. C) Serial axial CT sections through the lesion of the carotid artery with locations of the stenosis shown by red arrows. D) Serial axial PAI sections through the lesion of the carotid artery showing the NIRAPA signal.

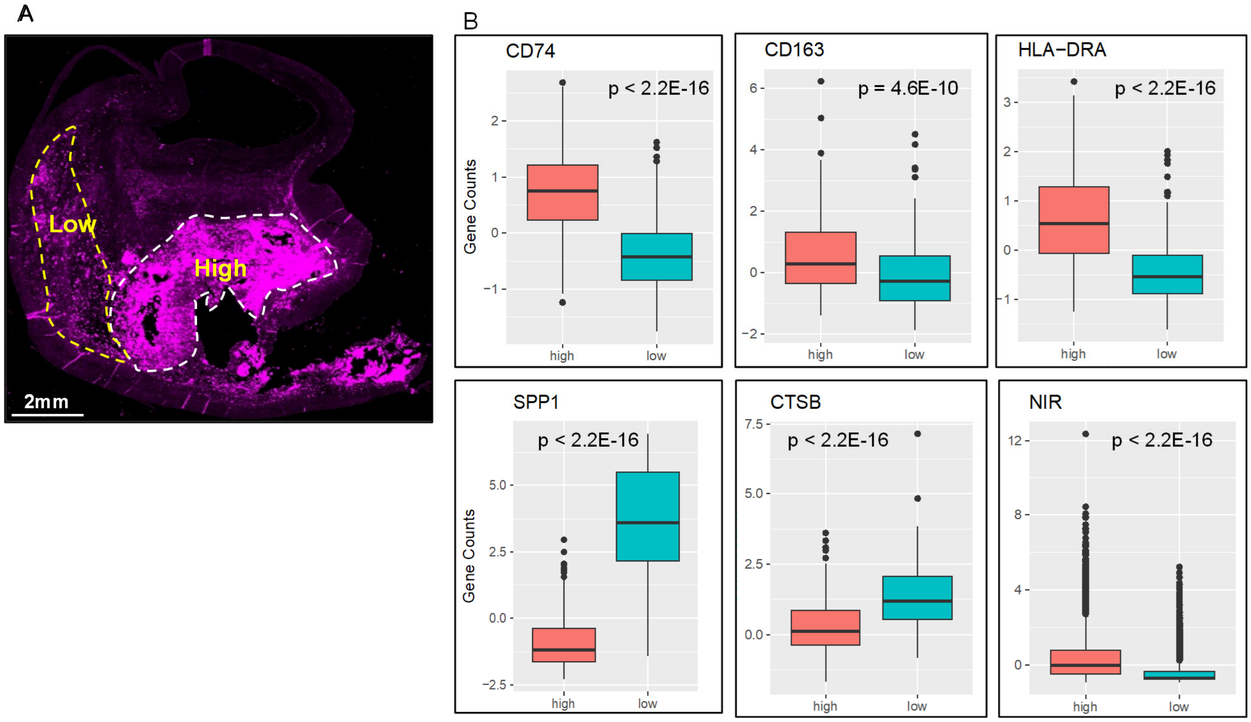


**Supplementary Fig. 3. Quantification and statistical comparison of the key gene counts corresponding to NIRAF signal regions based on Visium spatial transcriptomics with the high and low regions determined by NIRAF microscopic imaging.** A) Plaque regions with high and low NIRAF signal and B) *CD74*, *CD163*, *HLA-DRA*, *SPP1*, *CTSB* and NIRAF (NIR) signal level comparison between high and low signal regions and their p-values.

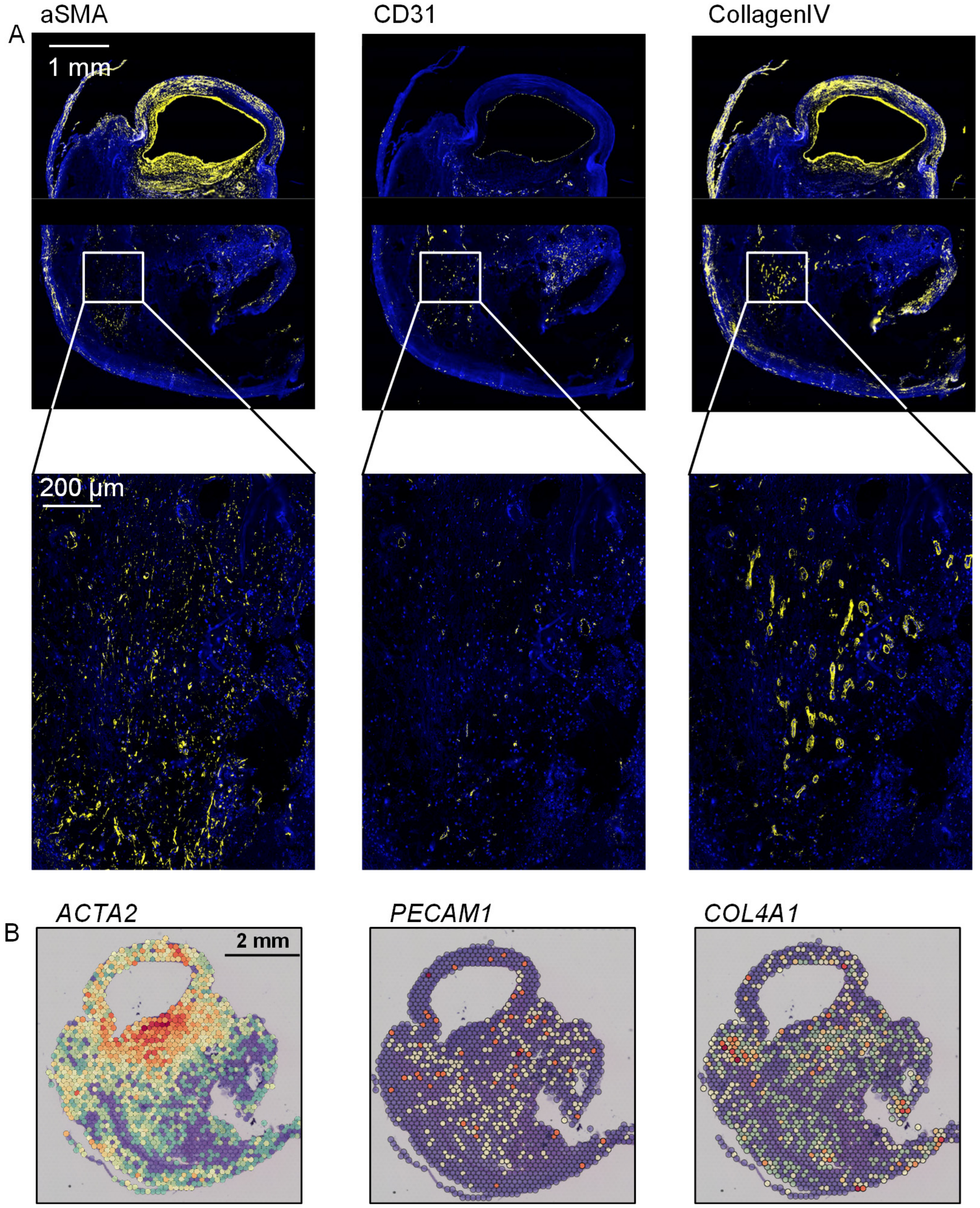


**Supplementary Fig. 4. Spatial correspondence of the transcriptomic and proteomic signals for protein and gene pairs.** Comparison of the A) proteomic and B) transcriptomic maps across the plaque for the following protein/gene pairs: αSMA protein and *ACTA2* gene, CD31 protein and *PECAM1* gene, and Collagen IV protein and *COL4A1* gene.

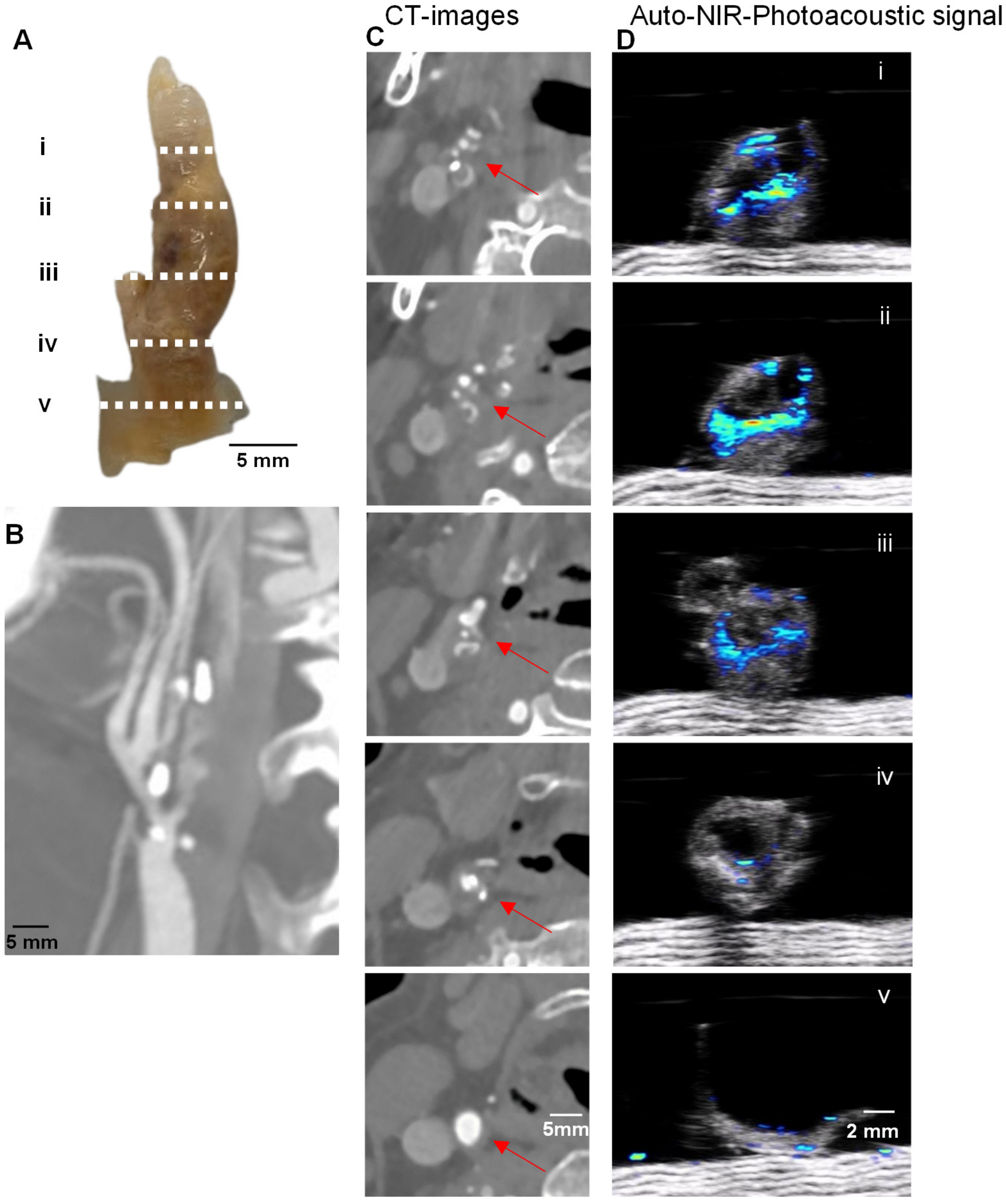


**Supplementary Fig. 5. Comparison of contrast CT images with PAI images using the NIRAPA signal to detect vulnerable plaque compositions of a vulnerable carotid artery.** A) Human carotid endarterectomy (CEA) specimen under white light. B) corresponding longitudinal contrast CT image shown underneath. The dashed lines on the longitudinal image represent the imaging locations of the axial images in columns B and C. C) Serial axial CT sections through the lesion of the carotid artery. Red arrows indicate location of stenosis. D) Serial axial PAI sections through the lesion of the carotid artery showing the NIRAPA signal.

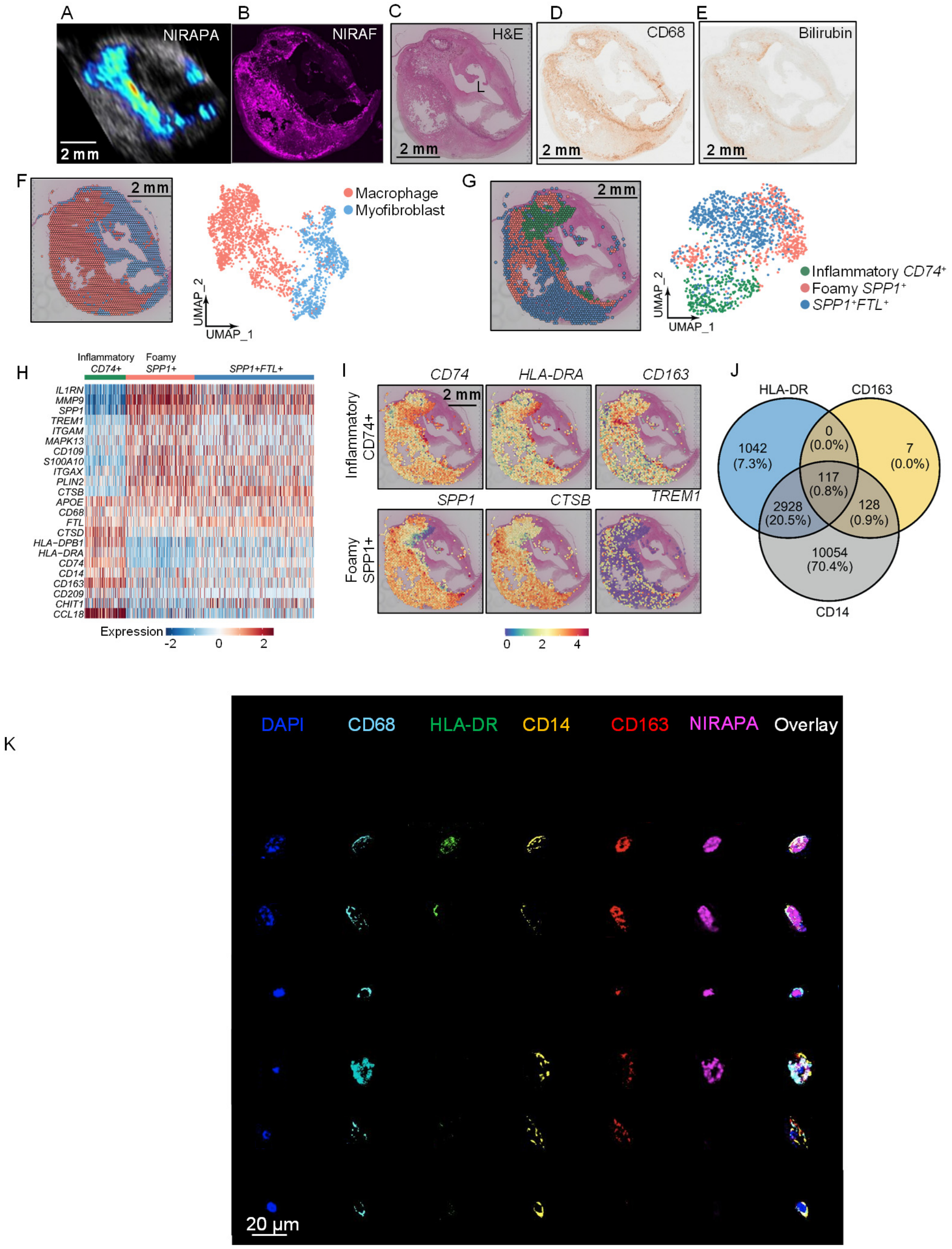


**Supplementary Fig. 6. Spatial transcriptomic and proteomic analysis of vulnerable plaque delineates spatially-dependent macrophage populations.** A-B) NIRAPA (A) and NIRAF (B) images of a carotid plaque cross section. C-E) Histological sections of the carotid endarterectomy (CEA) specimen stained with H&E, CD68 and bilirubin. F) Overlay and Uniform Manifold Approximation and Projection (UMAP) cluster projection of spatial transcriptomics on carotid plaque H&E. Based on their gene expression, clusters have been assigned to macrophage, myofibroblast and SMC intermediate cell types. G) Spatial deconvolution and UMAP of the macrophage cluster in CD74$^{+}$ and SPP1$^{+}$ regions and the spatial location on the H&E cross section. UMAP projection of macrophage high resolution subtype clustering shows CD74$^{+}$, SPP1$^{+}$, and *SPP1*$^{+}$*FTL*$^{+}$ populations. H) Heatmap of macrophage-specific gene signatures that differentiate the *CD74*$^{+}$, *SPP1*$^{+}$, and *SPP1+FTL+* macrophage subpopulations. I) Key genes differentiating inflammatory (*CD74*$^{+}$) and foamy (*SPP1*$^{+}$) macrophages and their spatial location on the CEA specimen. J) Venn diagram of macrophage markers based on single-cell identification on CODEX. K) CODEX imaging of individual cells within the plaque shown in A-E to validate the source of the NIRAPA signal. p value cutoff is 0.005. $Log_2FC$ cutoff is 2. L: Lumen.
